# Neurophysiology of Resilience in Juvenile Fibromyalgia

**DOI:** 10.1101/2024.06.05.24308376

**Authors:** Maria Suñol, Saül Pascual-Diaz, Jon Dudley, Michael Payne, Catherine Jackson, Han Tong, Tracy Ting, Susmita Kashikar-Zuck, Robert Coghill, Marina López-Solà

**Author notes:** Corresponding Authors: **Maria Suñol, PhD**, Address: Department of Medicine, School of Medicine and Health Sciences, University of Barcelona, Casanova, 143, Ala Nord, 5a planta, Barcelona, 08036, Spain. E-mail address, **Marina López-Solà, PhD**, Address: Department of Medicine, School of Medicine and Health Sciences, University of Barcelona, Casanova, 143, Ala Nord, 5a planta, Barcelona, 08036, Spain. This study was funded by Cincinnati Children’s Hospital Medical Center’s Trustee Grant Award and NIH/NIAMS Grants R01 AR074795 and P30 AR076316. Maria Suñol, PhD, is hired through the Ayuda para contratos Juan de la Cierva Formacion, funded by MICIU/AEI/10.13039/501100011033 and by the European Union NextGenerationEU/PRTR. Marina Lopez-Sola, PhD, is hired as part of the Serra Hunter Programme of the Generalitat de Catalunya. The authors have no conflict of interest to declare.

## Abstract

**Objective:** Juvenile fibromyalgia (JFM) is a chronic pain syndrome predominantly affecting adolescent girls. Resilience may be a protective factor in coping with pain, reducing affective burden, and promoting positive outlooks. Brain regions affected in JFM overlap with those linked to resilience, particularly in the default-mode network (DMN). We investigate the role of resilience on core somatic and affective symptoms in JFM and assess the neurophysiological substrates for the first time.

**Methods:** Forty-one girls with JFM and 40 pain-free adolescents completed a resting-state fMRI assessment and self-report questionnaires. We used clustering analyses to group JFM participants based on resilience, and principal component analyses to summarize core somatic and affective symptoms. We estimated whole-brain and within-DMN connectivity and assessed differences between higher and lower resilience JFM groups and compared their connectivity patterns to pain-free participants.

**Results:** The higher resilience JFM group had less affective (T=4.03; p<.001) but similar core somatic symptoms (T=1.05; p=.302) than the lower resilience JFM group. They had increased whole-brain (T’s>3.90, pFDR’s<.03) and within-DMN (T=2.20, p=.03) connectivity strength, and higher connectivity between DMN nodes and self-referential, regulatory, and reward-processing regions. Conversely, higher DMN-premotor connectivity was observed in the lower resilience group.

**Conclusion:** JFM participants with higher resilience were protected affectively but not in core somatic symptoms. Greater resilience was accompanied by higher signal integration within the DMN, a network central to internally oriented attention and flexible attention shifting. Crucially, the connectivity pattern in highly resilient patients resembled that of pain-free adolescents, which was not the case for the lower resilience group.

## 1. Introduction

Juvenile fibromyalgia (JFM) is a chronic pain syndrome that affects 2-6% of youth, primarily female adolescents during a crucial period of brain, body, and psychosocial development. JFM is characterized by widespread musculoskeletal pain, fatigue, and sleep and mood disturbances^1,2^. Up to 80% of patients experience symptoms into adulthood, leading to continued impairment^3^. In an 8-year follow-up study, JFM patients with worsening depressive symptoms experienced declining physical functioning, while those with stable or improving symptoms did not. This study highlights the importance of an affective component in explaining functional outcomes in JFM and suggests that a subgroup of patients may be at-risk for worsening emotional and physical functioning, while others demonstrate greater resilience^3^. Positive psychological factors such as optimism, sense of purpose, active coping, and pain acceptance have been linked to lower disease burden and improved functioning in adults with chronic pain^4,5^. In adolescents with chronic musculoskeletal pain, greater resilience has been preliminarily associated with reduced pain levels, physical disability, symptom severity, and suicidality, and with increased energy levels and health-related quality of life^6,7^. Psychological resilience may therefore be a protective factor against adverse outcomes in JFM and contribute to improved well-being and functioning over time.

JFM remains poorly understood from a pathophysiological perspective. Resting-state functional connectivity, a neuroimaging technique that measures synchronized activity between brain regions during rest, can offer insight into the intrinsic functional organization of the brain. Using this technique, we found that adolescent females with JFM had reduced cortico-cortical sensory integration and increased sensory-affective connectivity during rest^8^. Additionally, we found that volumetric gray matter changes in regions involved in emotional, self-referential, and language-related processing in patients were associated with lower well-being^9^. Thus, neurobiological alterations in JFM may extend beyond the nociceptive/somatosensory/broad sensory domains and involve impairments in emotional and self-referential processing areas.

Similarly, little is known about the neural correlates of resilience in youth, mainly due to heterogeneity in methods and in the definition and operationalization of resilience across studies. Nevertheless, recent reviews concur in suggesting a role of functional nodes of the default-mode network (DMN), such as the ventral prefrontal/orbitofrontal cortices and the precuneus, involved in self-referential and affective processing, and of subcortical regions such as the amygdala, hippocampus, and ventral striatum, -broadly involved in affective and stress-related responses, learning and memory-as linked with resilience in youth^10,11^. In addition, increased connectivity within DMN nodes and between DMN nodes and basal ganglia at rest has been associated with enhanced cognitive flexibility^12,13^. Cognitive flexibility is a key component of resilience, as it enables the shifting of attention away from negative or pain-related stimuli towards other tasks, facilitating adaptive coping strategies and maintaining daily functioning^14^. Nevertheless, most neuroimaging studies conceptualized resilience as the absence of clinical symptoms of psychopathology despite adversity. More comprehensive and nuanced approaches, such as measuring individual differences in coping strategies like “shifting” and “persisting” towards goals in response to adversity, offer an alternative perspective^15,16^. “Shifting” involves the ability to mentally reframe the meaning of a negative event, such as seeing a challenge as an opportunity, thus, it entails cognitive flexibility, whereas “persisting” involves the ability to maintain a positive attitude and continue striving towards a goal, even in the face of obstacles^15,16^. This approach recognizes resilience as not merely the absence of psychopathology but as a dynamic construct that encompasses a person’s capacity for positive adaptation and growth when facing adversity. This broader perspective can provide a more accurate and comprehensive understanding of the neural mechanisms associated with resilience.

Brain regions affected in JFM show overlap with areas associated with resilience in prior studies in youth, particularly involving DMN nodes^9–11,17^. DMN regions are particularly active during rest, when attention is not oriented towards an external object/task, but towards internally-oriented mentation^18,19^. This network plays a key role in shaping one’s sense of self and identity and has been extensively implicated in cognitive flexibility, underscoring its contribution in identity formation and adaptive thinking, particularly during the critical period of adolescence^12,13,20^. As shown, resilience holds promise for improving JFM outcomes and contributing to improved well-being and functioning over time^3,6,7^. However, are highly resilient JFM patients protected against core symptoms, such as symptom severity and widespread bodily pain? Or, is the effect more specifically associated with a protective affective profile? And, what are the neurophysiological substrates associated with high resilience in JFM? Is there a protective patient profile in terms of intrinsic brain organization? To answer these questions we studied, for the first time, differences between higher *vs.* lower resilience groups of JFM adolescent females on 1) core disease symptoms, 2) affective symptoms, and 3) brain functional connectivity. We hypothesize that the higher resilience group will exhibit less core disease-related and affective symptoms and increased resting-state functional connectivity strength in brain areas involved in affective and self-referential processing and in attention shifting, such as DMN regions. Studies such as this one may offer insights into the interplay of resilience, symptoms, and brain connectivity in JFM, shedding light on potential protective factors and avenues for intervention.

## 2. Methods

### 2.1 Participants

This study included 41 female adolescents diagnosed with JFM (16.6±1.05 years). We enrolled exclusively female participants because studies on clinical samples show that JFM is significantly more prevalent in girls^21,22^. In addition, 86% of eligible JFM participants from the parent clinical trial FIT-teens^23^ were cis-females. Thus, including other genders would have resulted in too small a representation given the possibility of relevant sex/gender differences in pain processing^24,25^ and brain developmental stage^26^. Such differences could have led to non-addressable confounding effects. In exploratory post-hoc analyses, we also used data of 40 pain-free female adolescents (16.3±.90 years). Inclusion criteria are detailed in **Supplement 1**.

Participants and their parent/legal guardians provided written informed consent/assent. The Cincinnati Children’s Hospital Medical Center’s Institutional Review Board approved the study protocol and consent forms (ID: 2017-7771). The study design, sample size calculations, and analysis plan were preregistered on the Open Science Framework (OSF) website (Registration DOI: 10.17605/OSF.IO/5CBZ6), in line with current best research practices.

### 2.2 Measures

Developmentally appropriate and validated self-report measures were used to assess sociodemographic and clinical characteristics. Resilience was measured with the 14-item Shift-and-Persist Scale, assessing how individuals cope with stress and adversity via two strategies: shifting and persisting, described in the introduction^15,16^. Core JFM symptom severity was assessed using the Pain and Symptom Assessment Tool^2,27^ which includes the Widespread Pain Index, in which participants report body areas where they had pain during the last three months and the Symptom Severity Checklist in which they indicate the severity of somatic symptoms (e.g., fatigue, headaches, dysautonomia, etc). Affective symptoms were assessed using the Children’s Depression Inventory^28^, which measures depressive symptoms in the past two weeks; the Screen for Child Anxiety-Related Disorders^29^, which screens for signs of anxiety disorders and provides five subscales that parallel the DSM-IV classification of anxiety disorders: general anxiety disorder, separation anxiety disorder, panic disorder, social phobia, and school phobia; and the Self-Compassion Scale^30^, which measures six components of self-compassion: Self-kindness, self-judgment, common humanity, isolation, mindfulness and over identification.

### 2.3 Imaging Data Acquisition, Preprocessing and Denoising

We collected resting-state BOLD functional magnetic resonance imaging and structural T1-weighted data with a Philips Ingenia 3.0-Tesla MR System (Philips Healthcare) equipped with a 32-channel head coil at the Cincinnati Children’s Hospital Medical Center. Imaging data were preprocessed and denoised using standard pipelines from the CONN Toolbox-20.b^31^ running on MATLAB-R2021a (Math Works Inc). Imaging data acquisition, preprocessing, and denoising are detailed in **Supplement 2**.

### 2.4 Data Analysis

#### 2.4.1 Hierarchical clustering of female adolescents with JFM based on resilience

We conducted an average linkage clustering analysis on the 41 JFM patients using normalized “shift” and “persist” scores from the Shift-and-Persist scale, a proxy for resilience. The silhouette test indicated two optimal clusters. This method resulted in one large, variable cluster with 37 patients and a smaller cluster with 4 patients scoring lowest in “shift” and “persist.” We then applied a three-cluster solution, yielding an 18-patient higher resilience cluster and two lower resilience clusters (19 and 4 patients), that we combined into a single lower resilience cluster of 23 patients. Cluster differences in “shift” and “persist” scores are shown in **Supplementary** Figure 1 and **Table 1**. We used χ2 and two-sample t-tests to compare demographic and clinical variables between higher and lower resilience groups. These analyses were performed with R software (R-project.org^32^). To confirm that the 4 JFM subjects with the lowest resilience, who formed a separate cluster in both the two-cluster and three-cluster solutions of the hierarchical clustering analyses, were not driving the behavioral/connectivity differences between the higher and lower resilience clusters, we replicated all analyses excluding them. The findings remained consistently unchanged.

**Table 1.** Differences in demographic and clinical variables between female adolescents with juvenile fibromyalgia and higher vs lower resilience.

|  | JFM Higher<br>Resilience (n = 18) | JFM Lower<br>Resilience (n = 23) | Statistics |  |
| --- | --- | --- | --- | --- |
| DEMOGRAPHIC VARIABLES | Mean ± SD | Mean ± SD | T / X<sup>2</sup> | p-value |
| Age (years) | 16.70 ± 1.02 | 16.47 ± 1.08 | .694 | .492 |
| Race (C / NC) | 17 / 1 | 20 / 3 | .643 | .423 |
| Yearly Household Income (1–7) | 5.44 ± 1.79 | 4.43 ± 1.93 | 1.71 | .094 |
| Education Level of the Primary<br>Caregiver (1–5) | 3.94 ± .80 | 3.96 ± .98 | -.042 | .966 |
| Education Level of the Secondary<br>Caregiver (1–5) | 3.82 ± 1.07 | 3.75 ± 1.07 | .208 | .836 |
| RESILIENCE | Mean ± SD | Mean ± SD | T | p-value |
| Shift | 12.94 ± 2.15 | 9.04 ± 2.27 | 5.59 | <.001 |
| Persist | 14.56 ± 1.38 | 9.43 ± 2.41 | 8.03 | <.001 |
| CORE JFM SYMPTOMS | Mean ± SD | Mean ± SD | T | p-value |
| Symptom Severity | 9.00 ± 1.61 | 8.70 ± 2.16 | .498 | .621 |
| Widespread Pain Index | 11.89 ± 3.14 | 10.57 ± 3.53 | 1.25 | .219 |
| AFFECTIVE SYMPTOMS | Mean ± SD | Mean ± SD | T | p-value |
| Child Depression Inventory | 12.61 ± 8.62 | 24.22 ± 8.70 | -4.26 | <.001 |
| SCARED: Panic Disorder | 8.91 ± 7.33 | 12.10 ± 5.79 | -1.35 | .187 |
| SCARED: GAD | 7.73 ± 6.10 | 13.38 ± 4.40 | -3.02 | .005 |
| SCARED: SAD | 3.64 ± 3.53 | 5.00 ± 3.79 | -.988 | .331 |
| SCARED: Social Phobia | 7.91 ± 4.95 | 7.95 ± 3.90 | -.027 | .979 |
| SCARED: School Phobia | 2.73 ± 2.57 | 4.33 ± 2.35 | -1.78 | .086 |
| SCS: Self Kindness | 3.00 ± 1.00 | 2.03 ± .54 | 4.00 | <.001 |
| SCS: Common Humanity | 2.82 ± .93 | 2.29 ± .81 | 1.94 | .060 |
| SCS: Mindfulness | 3.12 ± 1.11 | 2.54 ± .79 | 1.91 | .064 |
| SCS: Self Judgement | 3.41 ± 1.00 | 2.16 ± .68 | 4.69 | <.001 |
| SCS: Isolation | 3.49 ± 1.27 | 2.36 ± .79 | 3.48 | .001 |
| SCS: Over Identification | 3.67 ± 1.01 | 2.40 ± .73 | 4.65 | <.001 |

Note: **Yearly household income** is shown using a scale of 1–7, where 1 = <\$24,999; 2 = \$25,000 to \$49,999; 3 = \$50,000 to \$74,999; 4 = \$75,000 to \$99,999; 5 = \$100,000 to \$124,999; and 6 = \$125,000 to \$149,999; 7 > \$150,000. **Primary caregiver** refers to the person who is primarily responsible for meeting the child's daily needs (mother in 74 cases, father in 6 cases); **Secondary caregiver** refers to the person who also plays a significant role in the child's care and well-being but may not have the primary responsibility (father in 67 cases, mother in 6 cases, not assigned for 7 cases). **Caregiver education level** is shown using a scale of 1–5, where 1 = less than high school; 2 = high school/GED; 3 = partial college or trade school; 4 = college graduate; 5 = postgraduate degree. C: Caucasian; GAD: Generalized Anxiety Disorder; JFM: juvenile fibromyalgia; NC: Non-Caucasian; SAD: Separation Anxiety Disorder; SCARED: Screen for Child Anxiety Related Emotional Disorders; SCS: Self-Compassion Scale; SD: standard deviation.

#### 2.4.2 Differences in core JFM symptoms and affective symptoms between clusters of JFM patients based on resilience

We performed two principal component analyses (PCA) using varimax rotation and the Kaiser Criterion to select components with eigenvalues greater than 1. This approach helped us reduce dimensionality while identifying interpretable components that significantly explain the variance in the dataset. The PCAs focused on 1) core JFM symptoms (total scores of the Fibromyalgia Symptom Severity and Widespread Pain Index) and 2) affective symptoms (Child Depression Inventory score, subscales of the Screen for Child Anxiety-Related Disorders, and the Self-Compassion Scale). We computed the correlation between the principal components of the two PCAs to check for overlap, finding a non-significant negative correlation (r=-0.32, p=0.08), indicating a largely independent relationship, which supports our choice of performing separate PCAs for core and affective symptoms. We then used t-tests to compare the principal components of core and affective symptoms between JFM patient clusters based on resilience (i.e., JFM patients with lower *vs*. higher resilience). Analyses were conducted using Jamovi, built on R.

#### 2.4.3 Voxel-wise resting-state functional connectivity analysis

In first-level analyses, we computed whole-brain, voxel-wise resting-state functional connectivity using the Intrinsic Connectivity Contrast (ICC) implemented in the CONN toolbox. ICC is a measure of node centrality at each voxel that characterizes the connectivity strength by averaging the squared correlation coefficient values (r^2^) of a given voxel with all the other voxels in the brain^33^. In second-level analyses, we assessed differences in connectivity strength between JFM patients with higher *vs.* lower resilience using a two-sample t-test approach. The statistical threshold was false discovery rate (FDR) cluster-level corrected p<.05, with a voxel-level threshold of p<.001. As a complementary analysis, we computed ICC in 40 matched pain-free adolescent females and assessed differences between the clusters of JFM patients and pain-free adolescents.

#### 2.4.4 Post-hoc seed-based connectivity focusing on the default mode network (DMN)

ICC, as a measure of node centrality, does not provide information regarding what specific connections of such nodes are altered. Thus, we performed post-hoc seed-based analyses to locate the specific functional connectivity patterns that were altered, as recommended by ICC developers^33^. Since differences in whole-brain connectivity strength were mainly located in areas of the DMN, we selected four large cortical regions from the FSL Harvard-Oxford atlas that correspond to key nodes of the DMN: the medial prefrontal cortex, the right and left angular gyri, and the posterior cingulate cortex (PCC). We used these four regions of interest (ROIs) as seeds of interest in seed-based analyses. Next, we evaluated seed-based connectivity differences between JFM patients with higher *vs.* lower resilience using two-sample t-tests and a statistical threshold of FDR cluster-level corrected p<.05, with a voxel-level threshold of p<.001.

#### 2.4.5 Connectivity strength within the default mode network (DMN)

As complementary analyses, we compared connectivity strength within the DMN between JFM patients with higher *vs.* lower resilience. Unlike seed-based analyses, which explored connections between DMN regions and the entire brain, this approach assesses the network’s internal connectivity, providing insights into the intrinsic properties of the DMN. Using the CONN toolbox, we built a connectivity matrix per subject, with Z-scores representing connectivity between the 246 ROIs of the Brainnetome Atlas^34^. We used the Brainnetome atlas because it offers a finer-grained parcellation of the brain, enabling a more precise characterization of connectivity patterns within the DMN. We used a one-voxel dilated version of Yeo’s 7-Network atlas^35^ DMN mask to isolate DMN-specific connectivity. We computed the mean strength of connectivity within the DMN for each subject using in-house coding accessible at <u>github.com/neuroPENlab</u>. Last, we used two-sample t-tests in Jamovi to compare connectivity strength between groups, with statistical significance set at p<.05.

## 3. Results

### 3.1 Demographic and clinical variables

The clusters of JFM patients with higher *vs.* lower resilience did not differ in demographic variables (see **Table 1**). Differences in clinical variables will be discussed in the next section using a PCA approach and are also presented in **Table 1**. Differences between the two clusters of JFM patients and pain-free adolescents in demographic and clinical variables are presented in **Supplementary Tables 1 and 2**.

### 3.2 Differences in core JFM symptoms and affective symptoms between clusters of JFM patients based on resilience

The PCA of core JFM symptoms, including symptom severity and widespread pain, returned a single summary component. Likewise, the principal component analysis of affective symptoms including depressive, anxiety symptoms and self-compassion, returned a single summary component. Following the advice of Field^36^, we kept all factor loadings greater than 0.3, which is also the standard in Jamovi. Component loadings are presented in **Supplementary Table 3**.

The component summarizing core JFM symptoms did not differ between the JFM groups with lower *vs.* higher resilience (T=1.05; p=.302). In contrast, the groups differed in the component summarizing affective symptoms (T=4.03; p<.001, Cohen’s d=1.47, indicating a large effect size). Considering component loadings, this finding reflects that the higher resilience JFM group had less depressive and anxiety symptoms and higher self-compassion than the lower resilience group. For confirmatory purposes, we also assessed between-group differences for each of the variables included in the PCA. These findings are displayed in **Table 1**.

### 3.3 Vowel-wise resting-state functional connectivity analysis

JFM patients with higher resilience (*vs.* lower resilience) showed increased voxel-wise connectivity (i.e., connectivity strength) in the posterior cingulate cortex (PCC) (T=6.22, pFDR=.025, Cohen’s d=1.99); angular gyri (T=5.19, pFDR=.001, Cohen’s d=1.66; T=3.91, pFDR=.024, Cohen’s d=1.25); superior frontal (T=5.86, pFDR=.017, Cohen’s d=1.88; T=5.20, pFDR<.001, Cohen’s d=1.66) and inferior temporal gyri (T’s > 4.7, pFDR<.015, Cohen’s d>1.5) (see **Figure 1** and **Table 2**).

**Figure 1.**
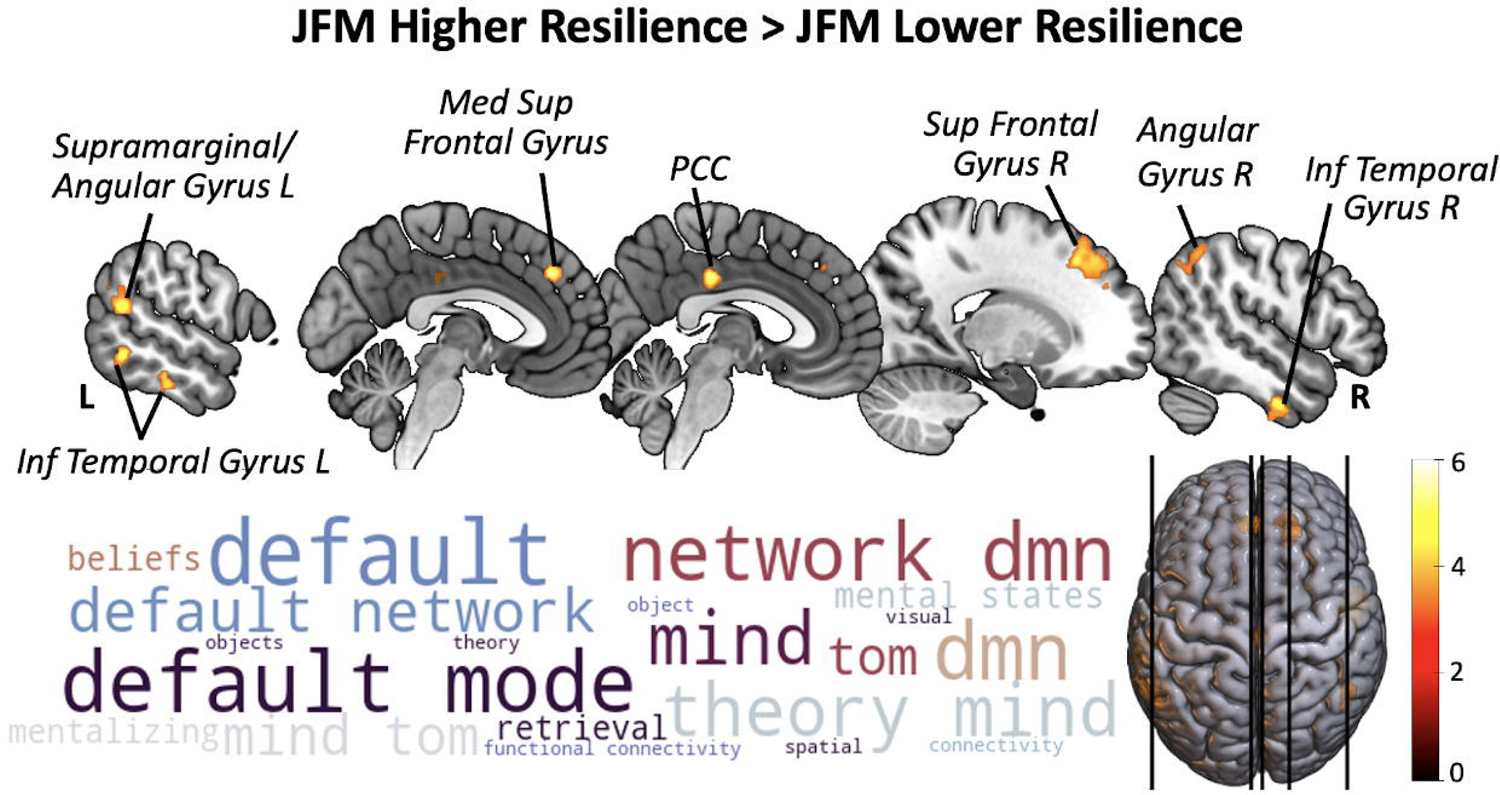
Voxel-based Connectivity Differences: JFM Higher vs Lower Resilience. **Top**: Clusters in warm colors represent global connectivity increases in the JFM subgroup with higher resilience compared to the lower resilience subgroup. These findings survive a voxel-level threshold of p<.001 and an FDR cluster-level corrected pFDR<.05. The color bar indicates T-values. Inf: Inferior; L: Left; Med: Medial; PCC: Posterior cingulate cortex; R: Right; Sup: Superior; **Bottom**: Display of the 20 functional terms most associated with the unthresholded t-map of this contrast based on the Neurosynth Database. Wordcloud was computed with Python. The size of the word is based on the correlation coefficient between each specific term and the connectivity t-map. The color of words is for aesthetic purposes. We found no significant differences in the JFM Higher Resilience < JFM Lower Resilience contrast.

**Table 2.**
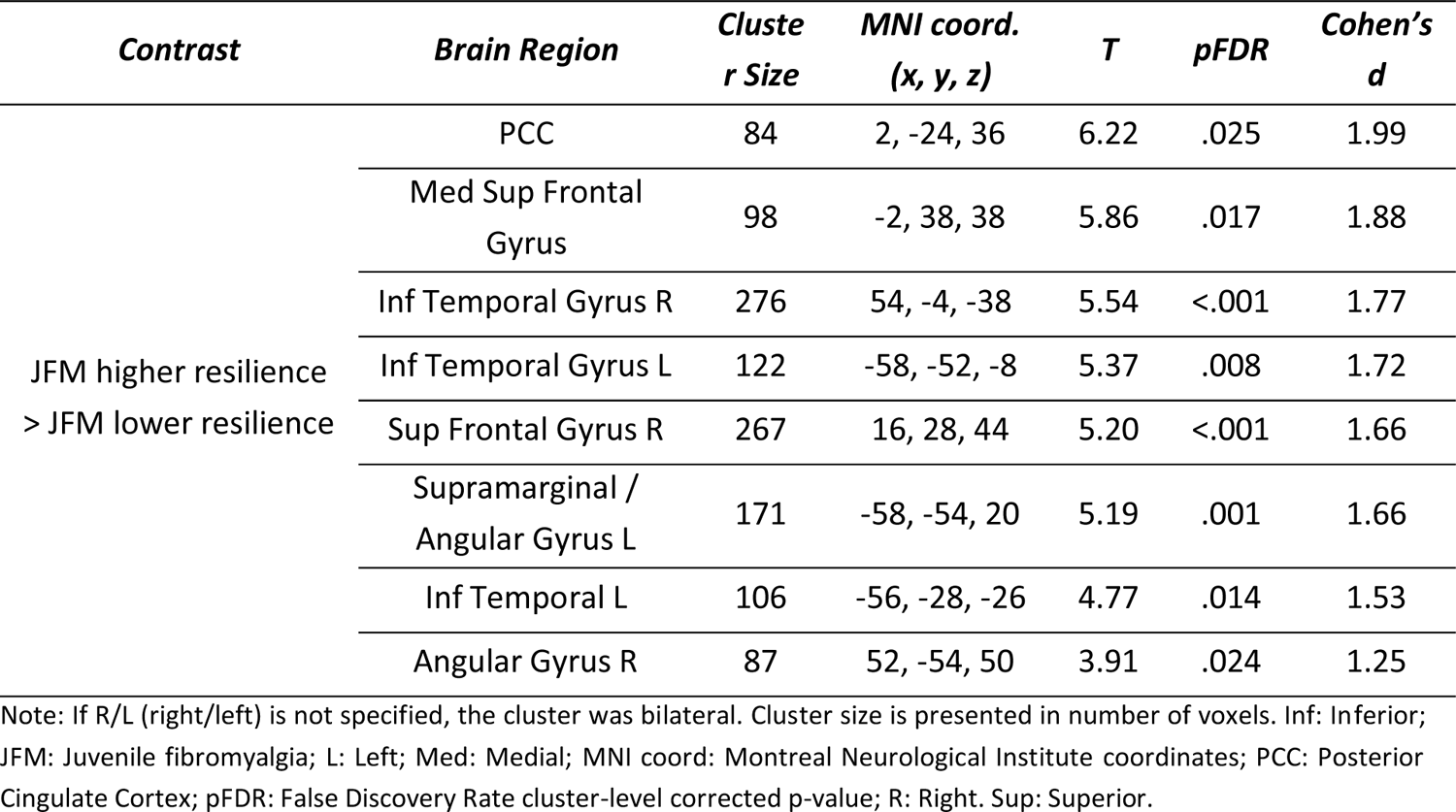
Differences in whole-brain, voxel-wise functional connectivity between female adolescents with JFM with lower vs higher resilience.

| *Contrast* | *Brain Region* | *Cluster Size* | *MNI coord.*<br>*(x, y, z)* | *T* | *pFDR* | *Cohen's d* |
| --- | --- | --- | --- | --- | --- | --- |
| JFM higher resilience<br>> JFM lower resilience | PCC | 84 | 2, -24, 36 | 6.22 | .025 | 1.99 |
|  | Med Sup Frontal Gyrus | 98 | -2, 38, 38 | 5.86 | .017 | 1.88 |
|  | Inf Temporal Gyrus R | 276 | 54, -4, -38 | 5.54 | <.001 | 1.77 |
|  | Inf Temporal Gyrus L | 122 | -58, -52, -8 | 5.37 | .008 | 1.72 |
|  | Sup Frontal Gyrus R | 267 | 16, 28, 44 | 5.20 | <.001 | 1.66 |
|  | Supramarginal / Angular Gyrus L | 171 | -58, -54, 20 | 5.19 | .001 | 1.66 |
|  | Inf Temporal L | 106 | -56, -28, -26 | 4.77 | .014 | 1.53 |
|  | Angular Gyrus R | 87 | 52, -54, 50 | 3.91 | .024 | 1.25 |

Note: If R/L (right/left) is not specified, the cluster was bilateral. Cluster size is presented in number of voxels. Inf: Inferior; JFM: Juvenile fibromyalgia; L: Left; Med: Medial; MNI coord: Montreal Neurological Institute coordinates; PCC: Posterior Cingulate Cortex; pFDR: False Discovery Rate cluster-level corrected p-value; R: Right. Sup: Superior.

Complementary post-hoc analyses including pain-free adolescents showed that, compared to these subjects, JFM patients with higher resilience had reduced connectivity in a single cluster of the paracentral lobule (PCL) (T=4.18; pFDR=.013, Cohen’s d=1.12). In contrast, compared to pain-free adolescents, JFM patients with lower resilience had a broader pattern of reduced connectivity encompassing sensorimotor, visual, attentional, and self-referential brain regions (T’s>4; pFDR’s<.04, Cohen’s d>1.02) (see **Figure 2** and **Table 3**).

**Figure 2.**
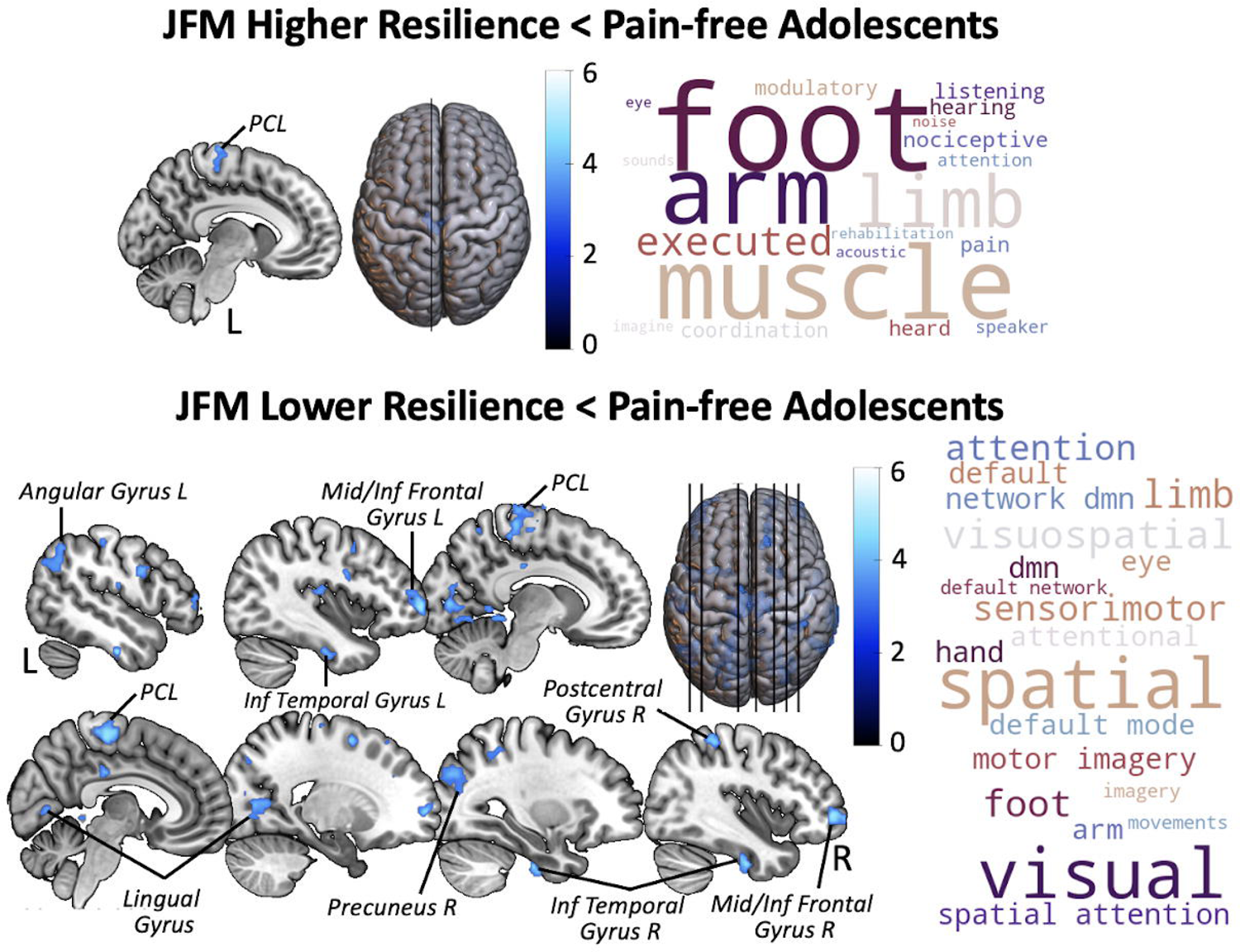
Voxel-based Connectivity Differences: JFM subgroups vs Pain-free Adolescents. **Top**: Clusters in blue represent global connectivity decreases in the JFM subgroup with higher resilience compared to pain-free adolescents. **Bottom**: Clusters in blue represent global connectivity decreases in the JFM subgroup with lower resilience compared to pain-free adolescents. All findings survive a voxel-level threshold of p<.001 and an FDR cluster-level corrected pFDR<.05. The color bar indicates T-values. Inf: Inferior; L: Left; Mid: Middle; PCL: Paracentral Lobule; R: Right. **On the right**, we display of the 20 functional terms most associated with the unthresholded t-maps of each contrast based on the Neurosynth Database. Wordcloud were computed with Python. The size of the word is based on the correlation coefficient between each specific term and the connectivity t-map. The color of words is for aesthetic purposes.

**Table 3.**
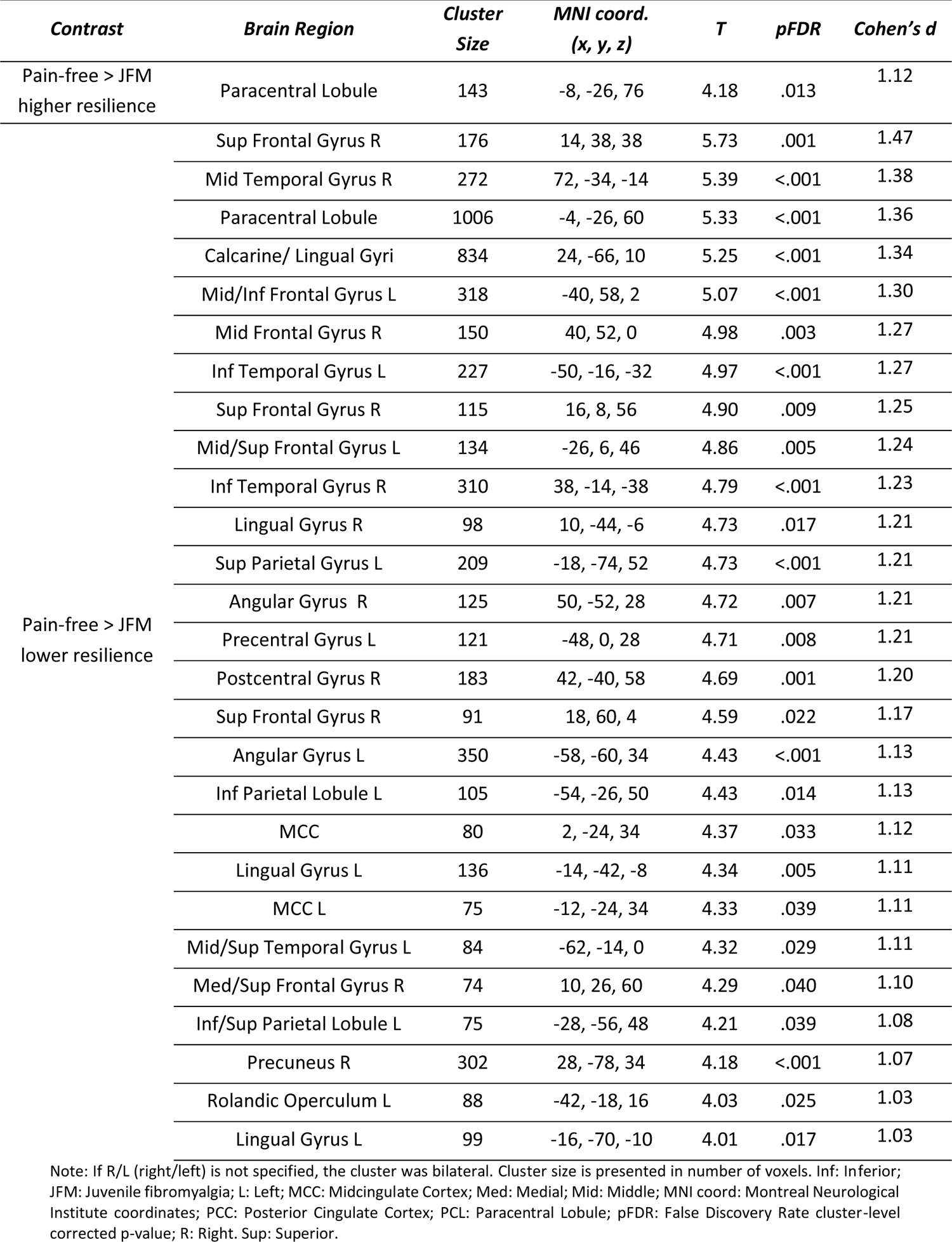
Differences in whole-brain, voxel-wise functional connectivity between clusters of JFM patients with lower and higher resilience compared to pain-free adolescents.

### 3.4 Post-hoc seed-based connectivity from key nodes of the default mode network (DMN)

Post-hoc analyses comparing seed-based connectivity from key nodes of the DMN between JFM patients with higher *vs.* lower resilience showed differential connectivity patterns. Subjects with higher resilience had increased connectivity between the PCC and the left angular and right inferior temporal gyri, whereas subjects with lower resilience had increased connectivity between the PCC and the right supplementary motor area (see **Figure 3** and **Supplementary Table 4**). JFM patients with higher resilience had increased connectivity between the angular gyri and the left caudate, expanding to the right orbitofrontal and medial superior frontal cortices at a trend level (pFDR=.057; p voxel-level<.001). Conversely, JFM patients with lower resilience had increased connectivity between the angular gyri and clusters in the cerebellum and premotor regions (see **Figure 3** and **Supplementary Table 4**). Medial prefrontal cortex connectivity did not differ between JFM subgroups.

**Figure 3.**
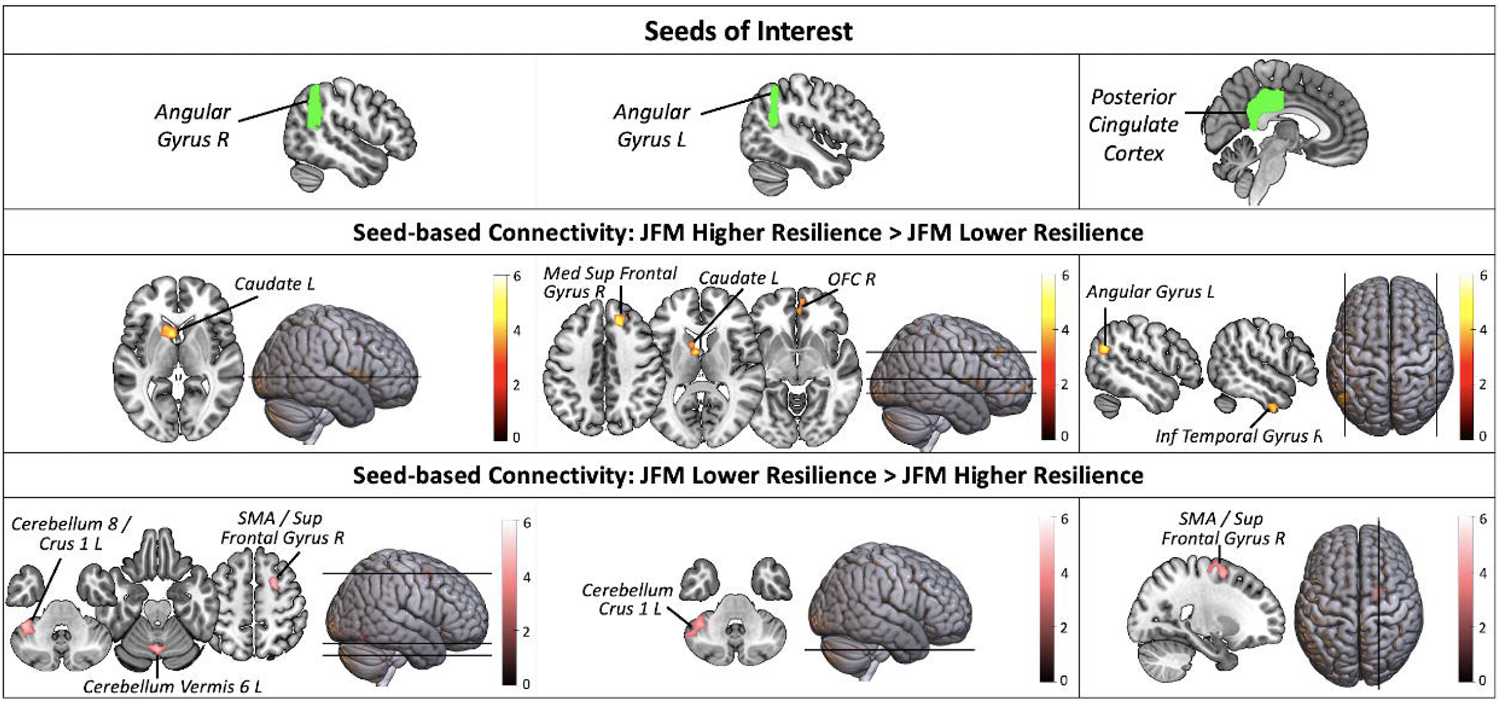
Seed-based connectivity differences between JFM subgroups from key nodes of the Default Mode Network. **Top**: Regions from the Harvard-Oxford Cortical Atlas used as seeds of interest in the seed-based analyses are displayed in green. The medial prefrontal cortex is not included here since its connectivity did not significantly differ between groups. **Middle**: Seed-based connectivity increases in JFM patients with higher resilience compared to the lower resilience JFM group are displayed in yellow. **Bottom**: Seed-based connectivity increases in JFM patients with lower resilience compared to the higher resilience JFM group are displayed in pink. All findings survive a voxel-level threshold of p<.001 and an FDR cluster-level corrected pFDR<.05, except the regions hyperconnected with the left angular in the higher (vs. lower) resilience group, which trended towards significance (pFDR’s=.057, p voxel-level <.001). The color bars indicate T-values. Inf: Inferior; Med: Medial; OFC: OrbitoFrontal Cortex; SMA: Supplementary Motor Area; Sup: Superior.

### 3.5 Connectivity strength within the default mode network (DMN)

In line with the rest of our findings, connectivity strength within the DMN was higher in JFM patients with higher resilience compared to those with lower resilience (T=2.20, p=.034). Connectivity strength within the DMN was also higher in pain-free adolescents compared to JFM patients with lower resilience (T=2.54, p=.014). Notably, we found no differences between pain-free adolescents and JFM patients with higher resilience (T<0.01, p=.998).

## 4. Discussion

A key finding of this investigation is that JFM patients with high resilience appear to be protected at the affective symptom level but not at the core somatic symptom level. They exhibited a pattern of greater signal integration during rest in regions of the DMN, a brain network that is important for self-related processing and attention shifting, that was not observed in the lower resilience group. Seed-based analyses revealed that higher resilience JFM patients had higher connectivity strength within the DMN during the resting state (compared with lower resilience JFM), and higher connectivity between DMN nodes and areas involved in affective, regulatory, self-referential, decision-making, and reward processing. In contrast, the lower resilience JFM group had increased connectivity between DMN nodes and motor areas. Last, we found that the connectivity strength pattern of higher resilience JFM patients both at the whole-brain level and specifically within the DMN resembled that of pain-free adolescents, which was not the case for the lower resilience JFM group.

Interestingly, we did not find that higher resilience was associated with reduced core JFM symptoms. This contradicts earlier preliminary findings showing that, in 28 adolescents with chronic musculoskeletal pain, resilience was negatively correlated with pain level, physical disability, and symptom severity^6^. The discrepancy may stem from different resilience measures; we used the Shift-and-Persist scale, which focuses on two strategies closely related to cognitive flexibility, whereas Gmuca et al.^6^ used the Connor-Davidson Resilience Scale 10-item (CD-RISC-10)^37^, which measures more general resilience traits such as competence, tenacity, and control in stressful situations. Studies in adult chronic pain have consistently reported associations between resilience and reduced pain and disability^4,5^. Thus, whether the impact of resilience on core symptoms emerges later in life after a longer exposure to chronic pain or is already present in adolescence remains to be elucidated. Conversely, and in agreement with previous research^6,7^, we found that higher resilience was associated with reduced suffering in JFM. Patients who were more resilient had higher emotional well-being, even if they were experiencing similar amounts of pain and disease severity. Our observation suggests that the first protection arising from a resilient attitude is on the “suffering” component associated with the disease, potentially via facilitating attention shifting and self-regulation processes, rather than through direct attenuation of bodily pain symptoms. Given the link between affective suffering and unfavorable JFM outcomes^3^, psychological interventions targeting resilience -and potentially specifically self-kindness-may not only alleviate current affective suffering but may also safeguard against future negative outcomes across psychological and physical domains. Promisingly, psychological interventions such as cognitive-behavioral therapy have shown effectiveness in enhancing coping skills, reducing catastrophizing, and improving self-efficacy and functioning in JFM^38^.

Our study shows, for the first time, that resilience was associated with different functional connectivity patterns in JFM patients. The higher resilience JFM group (*vs.* lower resilience) showed enhanced functional connectivity strength in areas of the DMN during rest, including the anterior/dorsal PCC -which extended to more posterior/ventral aspects of the PCC at an uncorrected level of p<.005, 20 voxels, which has shown to appropriately balance type I and type II error rates^39^ -, the angular gyri, and medial frontal regions. This network shows activation decreases during externally-oriented attention tasks, and activity increases during periods in which no particular external task is being attended, and therefore brain resources are allocated to internally-oriented self-relevant mentation^18,19^. There is also increasing evidence supporting the link between DMN and cognitive flexibility. Specifically, the DMN has been implicated in updating cognitive context and facilitating transitions between cognitive states^13,40^. Importantly, the PCC cluster peak fell in the anterior/dorsal aspect of the PCC, which is thought to be specifically involved in shifting attentional focus through influencing the metastability of the brain^41^. Previous studies show that adults with chronic pain have reduced cognitive flexibility, possibly as a product of threat monitoring and hypervigilance toward painful stimuli^42^. In line, studies have reported alterations of the DMN in people with chronic pain, specifically, decreased connectivity during rest, and enhanced connectivity between nodes of the DMN and the salience network^43–46^. Conversely, JFM patients with higher resilience displayed the opposite pattern, exhibiting increased DMN connectivity strength both at the whole-brain level and specifically within this network. This upregulation of signal integration within the DMN may reflect a brain network configuration associated with higher resilience. Such a configuration may be linked to an enhancement of cognitive flexibility, facilitating flexible disengagement from pain, suffering, or other potentially threatening stimuli, and directing attention towards more adaptive cognitive states and internally-oriented mentation in JFM patients who have developed higher resilience over time.

Compared to pain-free adolescents, both JFM groups showed connectivity reductions in the paracentral lobule, which may be a common feature of JFM more closely linked to somatosensory aspects of pain, as suggested by a recent study in this sample^8^. The higher resilience JFM group had no other connectivity differences compared to pain-free adolescents, which reinforces our interpretation that, in these subjects, the DMN connectivity strength is comparable to that of pain-free subjects. Conversely, the lower resilience JFM group had additional connectivity reductions beyond the somatosensory integration network, encompassing areas of the default-mode, visual, and frontoparietal networks. Preliminarily, this finding suggests that although somatosensory integration alterations are present regardless of resilience, this psychological characteristic may act as a protective factor to broader connectivity alterations.

Complementary analyses showed that the higher resilience JFM group had increased connectivity within the DMN and between nodes of the DMN and the left caudate, orbitofrontal cortex and right medial superior frontal cortex. These findings suggest that individuals with higher resilience may exhibit not only a more internally cohesive DMN but also stronger connections with brain areas that support cognitive control, emotional regulation, reward processing, and decision-making. In line, a previous study found that increased connectivity between DMN nodes and the basal ganglia at rest was associated with enhanced cognitive flexibility in pain-free adults^12^. In this context, our results may be interpreted as suggestive of a neural architecture associated with adaptive cognitive flexibility and self-regulation in individuals with higher resilience. The lower resilience JFM group exhibited reduced connectivity strength within the DMN paired with enhanced connectivity between DMN and premotor areas, involved in movement preparation and execution. Since the SMA is part of the cortico-striato-thalamo-cortical circuit, its hyperactivation during rest may shift motor circuits into an abnormal state of readiness^47^. Evidence suggests that, as age increases, SMA connectivity shifts from the DMN to attention and control networks. In younger individuals, the SMA-DMN coupling may enhance the translation of threat-related interoceptive signals and other threat-prone appraisals into motor commands^47,48^. Thus, JFM patients with lower resilience exhibit a DMN-premotor connectivity pattern akin to a delayed development, where the SMA may be triggered by self-relevant, threat-related signals, promoting an abnormal state of motor readiness, even at rest. In agreement, previous studies have reported similar increases in DMN-somatomotor connectivity to be associated with pain catastrophizing and anxiety symptoms in adults with chronic pain^46,49^. This state of motor readiness during rest may interfere with the ability to shift attention away from negative stimuli towards more adaptive responses, as evidenced by a previous study on 967 pain-free youth that found that youth with low cognitive flexibility (compared to the average) exhibited increased DMN-SMA connectivity^13^. Further research is warranted to examine the developmental processes involved with decreased resilience and determine the impact of psychological interventions in reducing these alterations before they become hard-wired in the brain in the transition into adulthood. Such research should consider the increasing evidence of social determinants of health, including socioeconomic status and adverse childhood experiences, which have lasting impacts on brain health and resilience^50^. Here, we observed a trend where the lower resilience group tended to have lower household income (T=1.71, p=.094), suggesting that early socioeconomic factors may influence resilience. This area is worthy of further investigation to better understand and address these contributing factors.

This study has important limitations. We enrolled cis-females, thus, our findings cannot be generalized to other genders. The rationale for this choice is detailed in the methods section. Future studies with adequately powered samples of cis-male and trans/non-binary individuals will allow examining between-sex/gender differences in JFM. Our sample had low representation of different ethnicities and participants with low socioeconomic status. Community-oriented research is crucial to overcome the predominance of white patients with medium/high socioeconomic status in research samples. Last, since this is the first study evaluating the connectivity correlates of resilience in youth with JFM, our findings should be replicated to determine their robustness.

In conclusion, our study sheds light on the significance of resilience in JFM, providing the first evidence of its impact on symptom presentation and brain functional connectivity early in the process of pain chronicity. Our results emphasize the clinical relevance of resilience in mitigating affective suffering and shaping neural networks, particularly the DMN. Given the association between affective suffering and unfavorable JFM outcomes^3^, psychological interventions targeting resilience may alleviate current affective suffering and safeguard against future symptom exacerbations and negative outcomes. Our findings suggest a link between resilience and brain network integrity in JFM. Notably, the lower resilience group displayed more extensive resting-state connectivity alterations, often resembling patterns observed in adult fibromyalgia. The fact that these connectivity alterations are observed in the context of JFM, a condition starting in youth, underscores the importance of early resilience-focused intervention, which may have the potential to reduce alterations before they become hard-wired, and to prevent the transition from juvenile to adult fibromyalgia.

## Data Availability

All data produced in the present study are available upon reasonable request to the authors

## Acknowledgements

This study was funded by Cincinnati Children’s Hospital Medical Center’s Trustee Grant Award and NIH/NIAMS Grants R01 AR074795 and P30 AR076316. Maria Suñol, PhD, is hired through the Ayuda para contratos Juan de la Cierva Formacion, funded by MICIU/AEI/10.13039/501100011033 and by the European Union NextGenerationEU/PRTR. Marina Lopez-Sola, PhD, is hired as part of the Serra Hunter Programme of the Generalitat de Catalunya. The authors gratefully thank Matt Lanier, Kaley Bridgewater, Kelsey Murphy, Brynne Williams, and Lacey Haas (Imaging Research Center, Department of Radiology, Cincinnati Children’s Hospital Medical Center) for their assistance in collecting MRI data.

