## Supplementary Material for "Neurophysiology of Resilience in Juvenile Fibromyalgia"

**Index:**

- **Supplement 1.** Inclusion and exclusion criteria
- **Supplement 2.** Imaging data acquisition, preprocessing, and denoising
- **Supplementary Figure 1.** Hierarchical Clustering of JFM participants based on resilience
- **Supplementary Table 1.** JFM higher resilience vs pain-free adolescents in demographic and clinical variables
- **Supplementary Table 2.** JFM lower resilience vs pain-free adolescents in demographic and clinical variables
- **Supplementary Table 3.** Principal component analyses of core JFM symptoms (left) and affective symptoms (right)
- **Supplementary Table 4.** Differences in seed-based functional connectivity between female adolescents with JFM with higher vs lower resilience

**Supplement 1.** Inclusion and exclusion criteria

Inclusion criteria for the JFM group included 1) JFM diagnosis from a pediatric rheumatologist or pain physician confirmed with 2010 American College of Rheumatology (ACR) criteria modified for pediatric use^1^. 2) Average pain intensity during the last week of 3 or greater out of 10, measured with the Brief Pain Inventory (BPI)^2^, and 3) Functional Disability Inventory (FDI)^3^ scores higher than 7 out of 60, indicating at least mild disability^4^.

Inclusion criteria for pain-free participants included 1) being physically and psychologically healthy (i.e., not diagnosed with chronic pain, psychiatric, neurological, or inflammatory disorders), and 2) an average pain score of 0 on a Numeric Rating Scale and an FDI score equal or below 7.

Subjects with contraindication to MRI scanning, developmental delay, major neurological or psychiatric disorders (except for JFM patients with mood, anxiety, or obsessive-compulsive disorders undergoing treatment), a positive pregnancy test, or taking opioid medication were not eligible. All participants included in the study were under no medication or under a stable medication regime for a minimum of 3 weeks prior to the MRI assessment.

**Supplement 2.** Imaging data acquisition, preprocessing, and denoising

We collected resting-state BOLD fMRI and structural T1-weighted data with a Philips Ingenia 3.0-Tesla MR System (Philips Healthcare, Best, Netherlands) equipped with a 32-channel head coil at the Cincinnati Children’s Hospital Medical Center. Resting-state BOLD fMRI data were collected using T2* weighted echo planar imaging sequence with a multiband sensitivity encoding (SENSE) technique with the following scan parameters: multiband acceleration factor = 4, TR = 650 ms, TE = 30 ms, field of view = 200 mm, flip angle = 53°, voxel size =2.5 x 2.5 x 3.5 mm, slice orientation = transverse (parallel to the orbitofrontal cortex line), slice thickness = 3.5 mm, number of slices = 40 (providing whole-brain coverage), number of volumes = 571, dummy scans = 12, and total scan duration = 6:13 min.

Imaging data were preprocessed and denoised using MATLAB-R2021a (Math Works Inc, Natick, MA) and the CONN Toolbox-20.b^5^ and following standard preprocessing steps: realignment and unwarping, outlier detection, segmentation and normalization of structural images, and normalization and smoothing of functional images. The first 12 volumes of the resting-state sequence were discarded to allow magnetization to reach equilibrium and the remaining volumes were realigned to the first one. Based on head-motion parameters estimated during realignment, volumes with framewise displacement >0.9mm or global BOLD signal changes >5 standard deviations were flagged as outliers. Realigned images were co-registered to the T1-weighted volume, which was normalized into standard Montreal Neurological Institute (MNI) space and segmented into gray matter, white matter, and cerebrospinal fluid compartments. Then, the same estimated non-linear transformation was applied to the functional images. Last, functional images were smoothed with a 6mm full width at half-maximum Gaussian kernel. In a subsequent temporal denoising step, noise components from white matter and cerebrospinal areas, characterized by the principal component-based “aCompCor” method implemented in the CONN toolbox^6^, were regressed out from BOLD timeseries together with subject-motion parameters, motion outliers, and a linear detrending term. Finally, we performed a band-pass filtering of 0.008Hz-0.09Hz. Images were inspected for artifacts at every step and quality assurance plots were visually examined.

**Supplementary Figure 1.** Hierarchical Clustering of JFM participants based on resilience

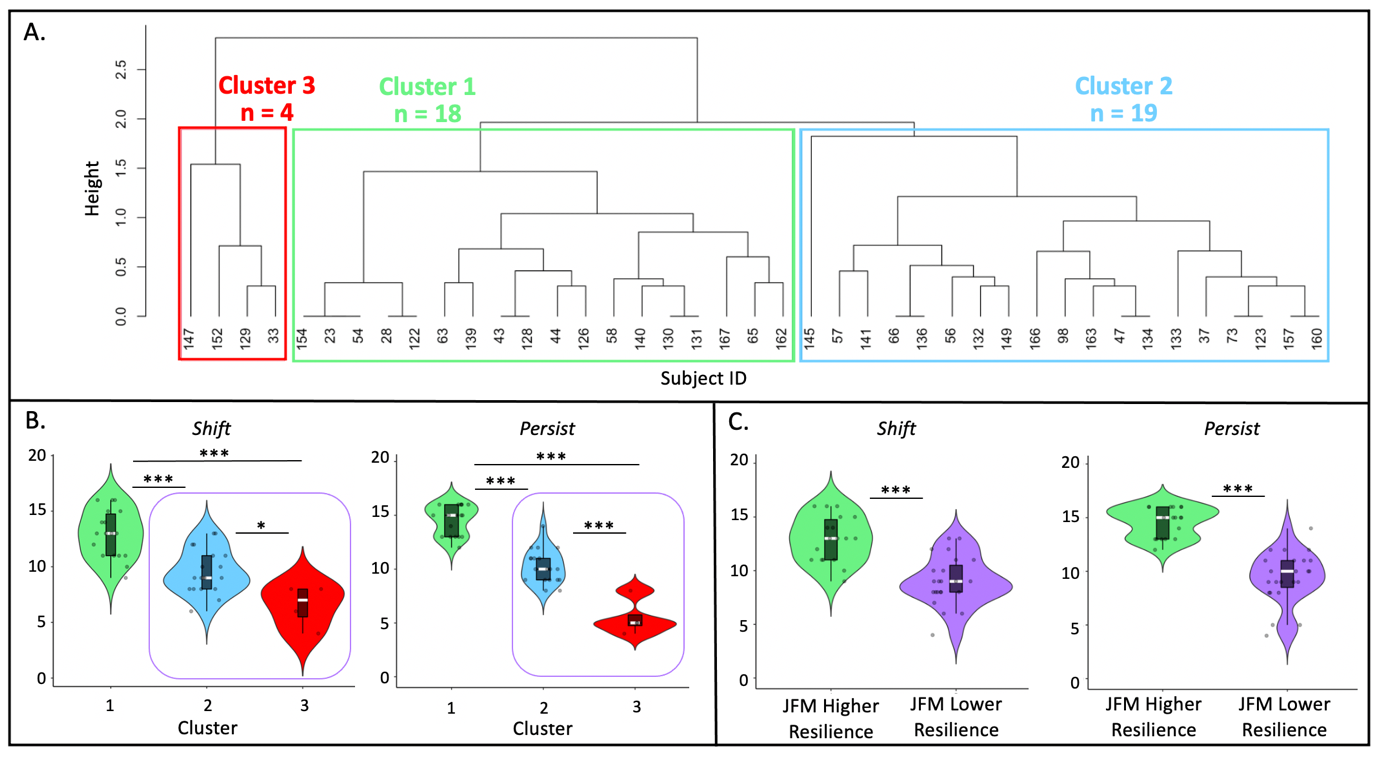

**A.** Hierarchical clustering of the 41 female adolescents with JFM based on the normalized “shift” and “persist” scores from the Shift-and-Persist scale, proxy measure of resilience. **B.** Violin plots of the “shift” and “persist” scores in each of the three clusters resulting from the hierarchical clustering analysis. Each box represents the 25th to 75th percentiles. White lines inside the boxes represent the median. Dark grey dots indicate individual scores. The purple square indicates the two clusters that were combined into the lower resilience JFM cluster for the final analyses. Connectivity and behavioral analyses were replicated excluding the 4 subjects with the lowest shift and persist values of cluster 3, and results remained unchanged, indicating that they were not driving between-clusters differences. **C.** Violin plots of the “shift” and “persist” scores after combining clusters 2 and 3 into the lower resilience JFM group. Cluster 1 constitutes the higher resilience JFM group. Each box represents the 25th to 75th percentiles. White lines inside the boxes represent the median. Dark grey dots indicate individual scores. *<.05; ***p < .001.

**Supplementary Table 1.** JFM higher resilience vs pain-free adolescents in demographic and clinical variables

|  | **JFM Higher Resilience (n = 18)** | | **Pain-free Adolescents (n = 40)** | **Statistics** | |
| --- | --- | --- | --- | --- | --- |
| **DEMOGRAPHICS** | | ***Mean ± SD*** | ***Mean ± SD*** | ***T*** | ***p-value*** |
| Age (years) | | 16.70 ± 1.02 | 16.31 ± .90 | 1.48 | .146 |
| Race (C / NC) | | 17 / 1 | 37 / 3 | .073 | .787 |
| Yearly Household Income (1–7) | | 5.44 ± 1.79 | 5.02 ± 1.98 | .768 | .446 |
| Education Level of the Primary Caregiver (1–5) | | 3.94 ± .80 | 4.10 ± .08 | -.661 | .512 |
| Education Level of the Secondary Caregiver (1–5) | | 3.82 ± 1.07 | 3.71± .90 | .406 | .687 |
| **RESILIENCE** | | ***Mean ± SD*** | ***Mean ± SD*** | ***T*** | ***p-value*** |
| Shift | | 12.94 ± 2.15 | 12.12 ± 2.93 | 1.05 | .297 |
| Persist | | 14.56 ± 1.38 | 14.41 ± 1.89 | 0.28 | .777 |
| **CORE JFM SYMPTOMS** | | ***Mean ± SD*** | ***Mean ± SD*** | ***T*** | ***p-value*** |
| Symptom Severity | | 9.00 ± 1.61 | 1.21 ± 1.45 | 17.03 | <.001 |
| Widespread Pain Index | | 11.89 ± 3.14 | .32 ±1.16 | 17.79 | <.001 |
| **AFFECTIVE SYMPTOMS** | | ***Mean ± SD*** | ***Mean ± SD*** | ***T*** | ***p-value*** |
| Child Depression Inventory | | 12.61 ± 8.62 | 5.09 ± 5.54 | 3.83 | <.001 |
| SCARED: Panic Disorder | | 8.91 ± 7.33 | 1.95 ± 2.66 | 3.77 | <.001 |
| SCARED: GAD | | 7.73 ± 6.10 | 3.47 ± 3.31 | 2.49 | .019 |
| SCARED: SAD | | 3.64 ± 3.53 | .74 ± 1.15 | 3.33 | .002 |
| SCARED: Social Phobia | | 7.91 ± 4.95 | 3.16 ± 3.55 | 3.06 | .005 |
| SCARED: School Phobia | | 2.73 ± 2.57 | .63 ± .76 | 3.34 | .002 |
| SCS: Self Kindness | | 3.00 ± 1.00 | 3.4 ± .96 | -1.41 | .165 |
| SCS: Common Humanity | | 2.82 ± .93 | 3.30 ± 1.15 | -1.53 | .132 |
| SCS: Mindfulness | | 3.12 ± 1.11 | 3.35 ± 1.07 | -.71 | .484 |
| SCS: Self Judgement | | 3.41 ± 1.00 | 3.67 ± .98 | -.90 | .327 |
| SCS: Isolation | | 3.49 ± 1.27 | 3.79 ± .94 | -.99 | .327 |
| SCS: Over Identification | | 3.67 ± 1.01 | 3.63 ± .94 | .12 | .903 |

Note: **Yearly household income** is shown using a scale of 1–7, where 1 = <$24,999; 2 = $25,000 to $49,999; 3 = $50,000to $74,999; 4 = $75,000 to $99,999; 5 = $100,000 to $124,999; and 6 = $125,000 to $149,999; 7 > $150,000. **Primary caregiver** refers to the person who is primarily responsible for meeting the child’s daily needs (mother in 74 cases, father in 6 cases); **Secondary caregiver** refers to the person who also plays a significant role in the child's care and well-being but may not have the primary responsibility (father in 67 cases, mother in 6 cases, not assigned for 7 cases). **Caregiver education level** is shown using a scale of 1–5, where 1 = less than high school; 2 = high school/GED;3 = partial college or trade school; 4 = college graduate; 5 = postgraduate degree. C: Caucasian; GAD: Generalized Anxiety Disorder; JFM: juvenile fibromyalgia; NC: Non-Caucasian; SAD: Separation Anxiety Disorder; SCARED: Screen for Child Anxiety Related Emotional Disorders; SCS: Self-Compassion Scale; SD: standard deviation.

**Supplementary Table 2.** JFM lower resilience vs pain-free adolescents in demographic and clinical variables

|  | **JFM Lower Resilience (n = 23)** | | **Pain-free Adolescents**  **(n = 40)** | **Statistics** | |
| --- | --- | --- | --- | --- | --- |
| **DEMOGRAPHICS** | | ***Mean ± SD*** | ***Mean ± SD*** | ***T*** | ***p-value*** |
| Age (years) | | 16.47 ± 1.08 | 16.31 ± .90 | .64 | .524 |
| Race (C / NC) | | 20 / 3 | 37 / 3 | .52 | .471 |
| Yearly Household Income (1–7) | | 4.43 ± 1.93 | 5.02 ± 1.98 | -1.15 | .255 |
| Education Level of the Primary Caregiver (1–5) | | 3.96 ± .98 | 4.10 ± .08 | -.61 | .541 |
| Education Level of the Secondary Caregiver (1–5) | | 3.75 ± 1.07 | 3.71± .90 | .15 | .882 |
| **RESILIENCE** | | ***Mean ± SD*** | ***Mean ± SD*** | ***T*** | ***p-value*** |
| Shift | | 9.04 ± 2.27 | 12.12 ± 2.93 | -4.24 | <.001 |
| Persist | | 9.43 ± 2.41 | 14.41 ± 1.89 | -8.72 | <.001 |
| **CORE JFM SYMPTOMS** | | ***Mean ± SD*** | ***Mean ± SD*** | ***T*** | ***p-value*** |
| Symptom Severity | | 8.70 ± 2.16 | 1.21 ± 1.45 | 14.73 | <.001 |
| Widespread Pain Index | | 10.57 ± 3.53 | .32 ±1.16 | 14.48 | <.001 |
| **AFFECTIVE SYMPTOMS** | | ***Mean ± SD*** | ***Mean ± SD*** | ***T*** | ***p-value*** |
| Child Depression Inventory | | 24.22 ± 8.70 | 5.09 ± 5.54 | 10.15 | <.001 |
| SCARED: Panic Disorder | | 12.10 ± 5.79 | 1.95 ± 2.66 | 7.00 | <.001 |
| SCARED: GAD | | 13.38 ± 4.40 | 3.47 ± 3.31 | 7.98 | <.001 |
| SCARED: SAD | | 5.00 ± 3.79 | .74 ± 1.15 | 4.70 | <.001 |
| SCARED: Social Phobia | | 7.95 ± 3.90 | 3.16 ± 3.55 | 4.05 | <.001 |
| SCARED: School Phobia | | 4.33 ± 2.35 | .63 ± .76 | 6.55 | <.001 |
| SCS: Self Kindness | | 2.03 ± .54 | 3.4 ± .96 | -6.21 | <.001 |
| SCS: Common Humanity | | 2.29 ± .81 | 3.30 ± 1.15 | -3.63 | <.001 |
| SCS: Mindfulness | | 2.54 ± .79 | 3.35 ± 1.07 | -3.06 | .003 |
| SCS: Self Judgement | | 2.16 ± .68 | 3.67 ± .98 | -6.27 | <.001 |
| SCS: Isolation | | 2.36 ± .79 | 3.79 ± .94 | -6.01 | <.001 |
| SCS: Over Identification | | 2.40 ± .73 | 3.63 ± .94 | -5.29 | <.001 |

Note: **Yearly household income** is shown using a scale of 1–7, where 1 = <$24,999; 2 = $25,000 to $49,999; 3 = $50,000to $74,999; 4 = $75,000 to $99,999; 5 = $100,000 to $124,999; and 6 = $125,000 to $149,999; 7 > $150,000. **Primary caregiver** refers to the person who is primarily responsible for meeting the child’s daily needs (mother in 74 cases, father in 6 cases); **Secondary caregiver** refers to the person who also plays a significant role in the child's care and well-being but may not have the primary responsibility (father in 67 cases, mother in 6 cases, not assigned for 7 cases). **Caregiver education level** is shown using a scale of 1–5, where 1 = less than high school; 2 = high school/GED;3 = partial college or trade school; 4 = college graduate; 5 = postgraduate degree. C: Caucasian; GAD: Generalized Anxiety Disorder; JFM: juvenile fibromyalgia; NC: Non-Caucasian; SAD: Separation Anxiety Disorder; SCARED: Screen for Child Anxiety Related Emotional Disorders; SCS: Self-Compassion Scale; SD: standard deviation.

**Supplementary Table 3.** Principal component analyses of core JFM symptoms (left) and affective symptoms (right)

| **PCA Core JFM Symptoms: 1 component** | | | **PCA Affective Symptoms: 1 component** | | |
| --- | --- | --- | --- | --- | --- |
| *Variables* | *Loadings* | *Uniq.* | *Variables* | *Loadings* | *Uniq.* |
| Symptom Severity | .834 | .304 | Child Depressive Inventory | -.791 | .374 |
| Widespread Pain Index | .834 | .304 | SCARED: Panic Disorder | -.645 | .585 |
|  |  |  | SCARED: GAD | -.931 | .133 |
|  |  |  | SCARED: SAD | -.627 | .607 |
|  |  |  | SCARED: Social Phobia | -.397 | .843 |
|  |  |  | SCARED: School Phobia | -.720 | .482 |
|  |  |  | SCS: Self Kindness | .702 | .507 |
|  |  |  | SCS: Common Humanity | .605 | .634 |
|  |  |  | SCS: Mindfulness | .602 | .637 |
|  |  |  | SCS: Self Judgement | .914 | .165 |
|  |  |  | SCS: Isolation | .892 | .205 |
|  |  |  | SCS: Over Identification | .857 | .265 |

Note: GAD: Generalized Anxiety Disorder; JFM: Juvenile Fibromyalgia; PCA: Principal Component Analysis; SAD: Separation Anxiety Disorder; SCARED: Screen for Child Anxiety Related Emotional Disorders; SCS: Self-Compassion Scale; Uniq.: Uniqueness.

**Supplementary Table 4.** Differences in seed-based functional connectivity between female adolescents with JFM with higher vs lower resilience

| *Contrast* | *Seed* | *Brain Region* | *Cluster Size* | *MNI coord.*  *(x, y, z)* | *T* | *pFDR* | *Cohen’s d* |
| --- | --- | --- | --- | --- | --- | --- | --- |
| JFM higher resilience > JFM lower resilience | PCC | Inf Temporal Gyrus R | 146 | 60, -4, -38 | 5.54 | .035 | 1.77 |
|  |  | Angular Gyrus L | 161 | -50, -62, 24 | 4.93 | .035 | 1.58 |
|  | Angular Gyrus L | Med Sup Frontal Gyrus R | 130 | 14, 36, 40 | 5.29 | .057^ | 1.69 |
|  |  | Caudate L | 147 | -4, 14, 4 | 4.67 | .057^ | 1.49 |
|  |  | OFC R | 119 | 6, 56, -6 | 3.81 | .057^ | 1.22 |
|  | Angular Gyrus R | Caudate L | 326 | -6, 12, 4 | 4.90 | .001 | 1.57 |
| JFM lower resilience > JFM higher resilience | PCC | SMA - Sup Frontal Gyrus R | 286 | 22, 4, 58 | 4.82 | .003 | 1.54 |
|  | Angular Gyrus L | Cerebellum Crus 1 L | 224 | -40, -48, -36 | 4.68 | .010 | 1.50 |
|  | Angular Gyrus R | Cerebellum Vermis 6 L | 226 | -2, -68, -24 | 5.25 | .004 | 1.68 |
|  |  | Cerebellum 8/Crus 1 L | 371 | -42, -42, -36 | 5.24 | <.001 | 1.68 |
|  |  | SMA - Sup Frontal Gyrus R | 137 | 22, 4, 56 | 4.94 | .029 | 1.58 |

Note: If R/L (right/left) is not specified, the cluster was bilateral. Cluster size is presented in number of voxels. Inf: Inferior; JFM: Juvenile fibromyalgia; L: Left; Med: Medial; MNI coord: Montreal Neurological Institute coordinates; OFC: Orbitofrontal Cortex; PCC: Posterior Cingulate Cortex; pFDR: False Discovery Rate cluster-level corrected p-value; R: Right. Sup: Superior; SMA: Supplementary Motor Area. ^ indicate results at a trend level of pFDR=.057 and p voxel-level<.001.
